# Educational interventions delivered to prescribing advisers to influence primary care prescribing: a very low-cost pragmatic randomised trial using routine data from OpenPrescribing.net

**DOI:** 10.1101/2024.01.05.24300907

**Authors:** Helen J Curtis, Brian MacKenna, Bhavana Reddy, Alex J Walker, Sebastian Bacon, Rafael Perera, Ben Goldacre

## Abstract

**Background:** NHS England issued commissioning guidance on 18 low-priority treatments which should not be routinely prescribed in primary care. We aimed to monitor the impact of an educational intervention delivered to regional prescribing advisors by senior pharmacists from NHS England on the primary care spend on low-priority items.

**Methods:** An opportunistic randomised, controlled parallel-group trial. Participants (clinical commissioning groups, CCGs) were randomised to intervention or control in a 1:1 ratio. The intervention group were invited to participate. The intervention was a one-off educational session. Our primary outcomes concerned the total prescribing of low-priority items in primary care. Secondary outcomes concerned the prescribing of specific low-priority items. We also measured the impact on information-seeking behaviour.

**Results:** 40 CCGs were randomised, 20 allocated to intervention, with 11 receiving the intervention. There was no significant impact on any prescribing outcomes. There was some possible evidence of increased engagement with data, in the form of CCG email alert sign-ups (p=0.077). No harms were detected.

**Conclusions:** A one-off intervention delivered to CCGs by NHS England did not significantly influence low-priority prescribing. This trial demonstrates how routine interventions planned to improve uptake or adherence to healthcare guidance can be delivered as low-cost randomised trials and how to robustly assess their effectiveness.

## Introduction

In 2017, NHS England issued commissioning guidance on 18 low priority treatments which should not be routinely prescribed in primary care (Table S1) [1]. We displayed the monthly trends in the population-adjusted spend on each of these items, for every practice and regional Clinical Commissioning Group (CCG) in England within the suite of tools available on OpenPrescribing.net - a free and openly accessible data service. Analyses carried out after 12 and 18 months of this information being openly available, indicated that this guidance had had little impact on the overall downward trend in expenditure across most of these medications [2,3].

At the time of this study, English primary care prescribing was funded by CCGs; these organisations have since transitioned into Integrated Care Boards (ICBs). CCGs took responsibility for promoting cost-effective and safe prescribing. In order to do this, most CCGs had (or shared) a Medicines Optimisation (MO) team, which can have significant influence on prescribing, e.g. implementing guidelines from National Institute for Health and Care Excellence (NICE) [4]. However, their activities vary, and wide variation in expenditure on low-priority treatments existed between CCGs. Our previous research found CCG membership to be a significant driver of variation in prescribing amongst their member practices, likely due to variation in CCG policies and activities [2,5].

As part of implementation work NHS England policy teams planned to visit CCGs with the highest spend on these low-priority products towards the end of 2018. The aim of the visits was to spread awareness of new commissioning guidance and collect feedback from CCGs around barriers to implementation. The visit consisted of a senior pharmacist giving a presentation about the aims of the guidance and featuring prescribing data for that CCG, and highlighting two drugs in particular, co-proxamol and dosulepin due to the strength of the recommendations associated with these two medications.

Since such policy interventions are commonly not evaluated, we aimed to conduct a very low cost randomised trial measuring its impact on the primary care prescribing. We measured the impact on the cost and rate of prescribing of low-priority treatments, and usage of OpenPrescribing by the CCGs and their constituent practices.

## Methods

### Trial Design

The protocol was released prospectively [6] and the trial was preregistered (ISRCTN31218900). It was designed as an agile evaluation using an opportunistic randomised controlled trial (RCT) to provide a robust assessment of impact for an intervention that was already planned by NHS England. This was a randomised, controlled parallel-group trial, with participants (CCGs) randomised to intervention or control in a 1:1 ratio. We were seeking to investigate superiority of the intervention over control. Interventions were conducted between October 2018 and January 2019. There were no substantial changes to methods after trial commencement; minor changes are described below.

### Participants

Participants were CCGs and staff therein. All 195 CCGs were eligible except those in which members of the study team were currently or recently employed (N=2). The invite letter, sent after randomisation, was addressed to the MO team [6].

### Randomisation

We identified the 40 CCGs with the greatest expenditure on 17 low-priority items (Table S1), adjusted for population size, during Jan-Jun 2018, the latest available six months of data. The 18th measure, on herbal remedies, was not available through OpenPrescribing at the time of randomisation due to complexity of identification of relevant products so was excluded from this step, but included in the intervention and in relevant outcomes. Prior to randomisation, we grouped CCGs known to have shared MO teams, and considered these as single units for the purpose of randomisation and intervention delivery. As this step reduced the number of eligible organisations, additional CCGs with the next-highest ranking for expenditure were included as necessary (see Results).

The 40 included organisations were randomised in a 1:1 ratio to receive the intervention or no intervention, implemented in software. Allocated CCGs were invited to participate. Participants were not informed that they were in a trial. The control group was not contacted. Outcome measurement and statistical analysis were performed using pre-prepared scripts in the study repository [7], so it was not necessary for researchers to be blinded to allocation. It was not possible for the NHS England staff delivering the intervention to be blinded.

### Interventions

The intervention was a single education session delivered in-person to each CCG individually by a senior NHS England representative (BMK/BR) at a location of each CCG’s choice. Each CCG/MO team was free to determine which staff members attended the intervention, which may have included staff outside of the MO team. The control group received no intervention.

Delivery was facilitated with the use of Microsoft Powerpoint slides and interactive demonstration with the OpenPrescribing.net website and the sessions lasted 1-2 hours. The presentation and outline notes for the presenter are available in the Supplementary Information and included:

- Standard presentation of rationale for guidance;
- Presentation of the organisations prescribing data compared to their peers
- This was presented using OpenPrescribing.net and information was provided on how to sign up for further automated feedback;
- Tailored presentation of data on two items included for safety purposes (co-proxamol and dosulepin);
- Tailored identification of “Top 3 areas for improvement” for that CCG;
- Sharing of best practice resources and other implementation tools;
- Time for reflection and a chance to ask presenters questions on any aspect of the guidance.

Interventions were planned to be carried out between October and December 2018. This was later extended to include January 2019 in order to schedule more interventions with limited staff availability. The planned outcome measures did not need to be altered to take this into account.

### Outcomes

Our primary objective was to determine whether receipt of a structured educated session at a CCG has any impact on prescribing behaviour in primary care, primarily based upon (P1) the cost of prescribing all 18 low-priority per 1000 registered population, over a six month follow-up period (April-September 2019) compared to baseline (April-September 2018). To support this we also measure (P2) the number of items prescribed per 1000 registered population, to account for any changes in medicine prices.

Our secondary objective was to assess the impact on specific low-priority items. Outcomes were (S1) the top three low-priority measures with the greatest spend for each CCG (this varies by CCG), measured as total spend across these items per 1000 population; and (S2) co-proxamol and (S3) dosulepin items prescribed across the same six-month follow-up period compared to baseline, per 1000 population.

To assess engagement with prescribing data we measured the impact on interaction with the OpenPrescribing.net website, firstly in terms of website page views covering low-priority measures, one month after versus one month before the intervention. This was conducted separately for CCG pages (E1) and practice pages grouped up to CCGs (E2). Secondly, we measured new registrations to OpenPrescribing’s monthly CCG (E3) and practice (E4) email alert service within 3 months of the intervention.

We planned to measure the change in CCGs incorporating the issue into their annual workplan. This manual data collection was not carried out so this outcome was not assessed. NHS England also collected qualitative feedback on barriers to implementation of the guidance but this was outside the scope of this evaluation.

We added a small number of additional analyses which were not pre-specified, as indicated in the text.

### Sample Size

The sample size was limited to 20 (in the intervention group) to ensure sufficient time and resources available to deliver the intervention within a 3-4 month period. It was not possible for participants to withdraw from analysis after allocation.

We estimated that we had 80% power to detect a difference of £31 per 1000 population (alpha=0.05) between the intervention and control group, based on the mean baseline measurement for the 40 eligible CCGs: £224 per 1000 population per month (SD 34). Summing over the six month baseline period to compare with outcome measures, this equates to a difference in total spend of £186 per 1000 and a mean of £1,344 per 1000 population. The median monthly spend at baseline across all CCGs was £150 per 1000 (75th percentile £174), therefore a decrease of £31 per 1000 population per month to a mean of £193 would represent a relatively small change in rankings. For the best CCG out of the 40 eligible (£178), a reduction of £31 per 1000 population per month would bring it to a ranking slightly below the median.

### Data Collection

Practice-level prescribing data (aggregated up to CCGs) was obtained from national datasets published monthly by NHS Digital, with approximately two months’ lag time [8]. Practice membership of CCGs and registered population sizes are also published by NHS Digital. This data is routinely compiled and loaded into our OpenPrescribing database in Google BigQuery. Baseline measurement of low priority prescribing for eligibility assessment (alongside allocation) was carried out using a Python script [7].

For engagement outcomes, we extracted data on “Unique Pageviews” (separate browsing sessions) for CCG dashboards from Google Analytics, for all eligible CCGs. We also counted the number of email alerts active for each CCG and practice from the OpenPrescribing email system.

Follow-up data was subject to analysis as pre-defined [6]. Minor changes made to the methods are described in Supplementary Note 1. Website access data was collected via Google’s Analysis services (users are not identifiable). Email alert sign-up data was downloaded from archives held within OpenPrescribing (email recipients are not identifiable).

### Statistical methods

#### Prescribing outcomes (P1-2, S1-3)

Measure values were calculated for each CCG by summing cost or items across the six-month baseline and follow-up period, averaging the total monthly CCG population, then calculating proportions (cost or items prescribed per 1000 population).

Outcomes were compared using regression models, with baseline value and intervention group as dependent variables. We initially planned to include the presence or absence of a workplan on low-priority prescribing reduction at baseline as a dependent variable; however survey results (NHS England) indicated that all CCGs who responded but one had implemented some or all of the recommendations and we did not include this in the pre-written analysis. We report p-values and 95% confidence intervals.

Missing data: none expected.

#### Engagement outcomes

Page views (E1-2): Analysis of total page views in the follow-up period (one month) used a regression model to compare intervention with control, with baseline (one month) page views as a co-variable.

Missing data: engagement data gathered from Google Analytics is expected to be complete except where users of the website have installed tools to prevent their activity being collected. This is likely to be uncommon, and equally distributed between intervention and control groups.

Email alert sign-ups (E3-4): Analysis of total email alert sign-ups within 3 months used a regression model, with the CCG registered population (as a proxy for staff numbers) and total number of sign-ups prior to the intervention as co-variables.

Missing data: none expected.

## Results

### Participant flow

Participant flow is summarised in Figure 1. Forty CCGs with the greatest expenditure on low-priority items were identified (after taking into account known joint working of MO teams); 20 were allocated to the control and 20 to intervention. Ten visits were conducted; however, due to shared MO teams, this covered 11 CCGs from the intervention group as well as six other non-allocated CCGs. For one CCG which accepted the invitation, a visit was not successfully scheduled. The remaining eight intervention CCGs did not respond to the invitation. All allocated participants were included in the intervention group for analysis.

**Figure 1.**
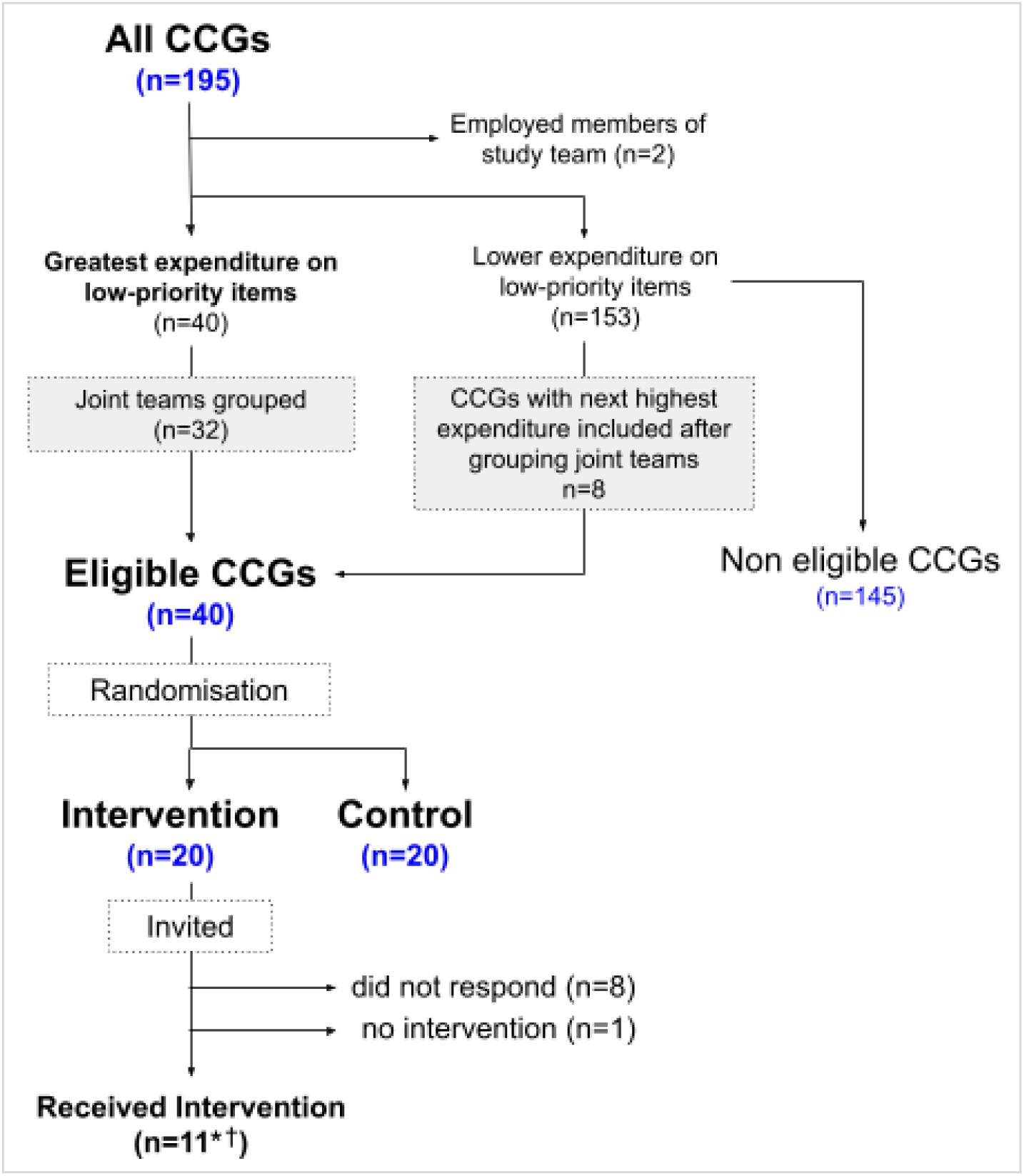
participant flow. *The intervention was delivered to CCGs with joint teams covering 6 additional CCGs from the “non eligible” group. ^†^two allocated CCGs were present at one intervention together, meaning 10 individual visits were conducted. CCG = Clinical Commissioning Group.

Randomisation was carried out and invitations sent on 26th Sep 2018, and visits were conducted between 26th Oct 2018 and 30th Jan 2019. Seven of the interventions were delivered to three or more attendees, two of which had ten or more attendees. The trial ran to completion as planned.

### Baseline Characteristics

The intervention group covered slightly larger populations, on average, than the control group (333.4k±154 vs 256.7k±119; Table 1). Practices in the intervention group spent slightly less on low-priority items than the control group during the baseline period (April-September 2018, £1,158±146.5 per 1000 population vs £1,289±224.3; Table 1) with slightly fewer items prescribed per 1000 population (30.6±10.6 vs 36.1±11.8). The proportion of patients over 65 was similar in both groups; this is a factor which has a strong association with spending on low-priority items [2].

**Table 1.** Baseline characteristics for intervention and control group.

|  | CCGs | Population (thousands) |  | Percent over 65 |  | Items per 1000 population |  | Spend per 1000 population |  |
| --- | --- | --- | --- | --- | --- | --- | --- | --- | --- |
|  | count | mean | std | mean | std | mean | std | mean | std |
| **intervention** | 20 | 333.4 | 154.0 | 19.1 | 4.0 | 30.6 | 10.6 | £1,158 | £146.5 |
| **control** | 20 | 256.7 | 119.9 | 20.2 | 4.4 | 36.1 | 11.8 | £1,289 | £224.3 |

*Population is the total registered population across the practices in each CCG. This and the percent over 65 were measured as of the end of the baseline period (April-September 2018) and prescribing figures were totalled across the six month baseline period. The baseline spend is lower than that from the power calculation due to the inclusion of additional CCGs with lower expenditure after grouping joint teams. std=Standard Deviation.*

### Primary Prescribing Outcomes

For our primary outcome (P1), the cost of all low-priority items per 1000 population in the follow-up period (April-September 2019), the spend was £959.0 for the intervention group and £1070.0 for the control group (Table 2); after taking into account baseline levels (£1,158.3 and £1,288.8 respectively), there was no difference between the two groups (p=0.695). Neither was there a significant difference when measuring items prescribed per 1000 population (P2), after adjusting for baseline level (24.9 vs 29.1, p=0.971, Table 2).

**Table 2.** Prescribing outcomes.

| All 18 “low-priority” treatments combined |  |  |  |  |  |  |  |  |  |  |
| --- | --- | --- | --- | --- | --- | --- | --- | --- | --- | --- |
| allocation | count | P1. Cost per 1000 population |  |  |  |  |  |  |  |  |
|  |  | Baseline |  | Follow-up |  | Change | Effect size | p value | 95% CI |  |
|  |  | mean | std | mean | std |  |  |  | lower | upper |
| control | 20 | £1,288.8 | £224.3 | £1,070.0 | £201.0 | -£218.8 |  |  |  |  |
| intervention | 20 | £1,158.3 | £146.5 | £959.0 | £152.4 | -£199.4 | -14.9 | 0.695 | -91.4 | 61.6 |
| P2. Items per 1000 population |  |  |  |  |  |  |  |  |  |  |
| control | 20 | 36.1 | 11.8 | 29.1 | 9.7 | -7.0 |  |  |  |  |
| intervention | 20 | 30.6 | 10.6 | 24.9 | 9.0 | -5.8 | -0.048 | 0.971 | -2.73 | 2.64 |
| S1. Top 3 pre-specified “low-priority” treatments per CCG<br>Cost per 1000 population |  |  |  |  |  |  |  |  |  |  |
| allocation | count | Baseline |  | Follow-up |  | Change | Effect size | p value | 95% CI |  |
|  |  | mean | std | mean | std |  |  |  | lower | upper |
| control | 20 | £866.8 | £225.7 | £664.1 | £206.8 | -£202.8 |  |  |  |  |
| intervention | 20 | £756.7 | £159.7 | £592.2 | £157.8 | -£164.5 | 15.9 | 0.629 | -50.2 | 82.0 |
| S2: Co-proxamol items per 1000 population |  |  |  |  |  |  |  |  |  |  |
| allocation | count | Baseline |  | Follow-up |  | Change | Effect size | p value | 95% CI |  |
|  |  | mean | std | mean | std |  |  |  | lower | upper |
| control | 20 | 0.295 | 0.160 | 0.199 | 0.123 | -0.096 |  |  |  |  |
| intervention | 20 | 0.262 | 0.232 | 0.176 | 0.203 | -0.086 | 0.003 | 0.894 | -0.040 | 0.046 |
| S3: Dosulepin items per 1000 population |  |  |  |  |  |  |  |  |  |  |
| control | 20 | 7.113 | 2.774 | 6.058 | 2.428 | -1.055 |  |  |  |  |
| intervention | 20 | 6.465 | 3.387 | 5.476 | 2.998 | -0.989 | -0.024 | 0.897 | -0.390 | 0.343 |

Primary outcomes P1 and P2: Total cost and number of items all 18 low-priority prescribed per 1000 population in the intervention versus control groups. Secondary outcomes S1-S3: Total cost of the top three low-priority items for each CCG (varies by CCG), and the number of co-proxamol and dosulepin items prescribed, per 1000 population in the intervention versus control groups.

We performed a sensitivity analysis (not pre-specified) where those CCGs which did not receive the intervention (n=9) were grouped into the control group. There remained no significant difference in either cost or items prescribed (p=0.39 and 0.73 respectively, Table S2).

### Secondary Prescribing Outcomes

For our secondary prescribing outcome S1, the cost of the top three low-priority items tailored for each CCG, there was no significant difference between intervention and control during the six-month follow-up period (£592.2 vs £664.1 per 1000 population respectively, p=0.629, Table 2). Neither was there a significant difference in co-proxamol (S2) and dosulepin (S3) items prescribed per 1000 population (p=0.894 and 0.897 respectively,Table 2).

### Impact on Engagement

For our primary engagement outcome E1, the mean number of page views over one month on CCG pages showing low-priority measures during follow-up was 0.65 for the intervention group (0.65 baseline) and 0.40 for the control group (0.10 baseline; Table 3); the regression model indicated there was no difference between the two groups (p=0.779). However, total page view counts were very low (2-13). Similarly, there was no significant difference in the pages viewed for practices, grouped up to their parent CCGs (2.20 to 1.00 vs 0.45 to 0.60, Table 2; p=0.517).

**Table 3.** Engagement outcomes.

| Page views of low-priority measures |  |  |  |  |  |  |  |  |  |  |  |  |
| --- | --- | --- | --- | --- | --- | --- | --- | --- | --- | --- | --- | --- |
| group | count | E1. CCG Pages |  |  |  |  |  |  |  |  |  |  |
|  |  | Before |  |  | After |  |  |  | Effect size | p value | 95% CI |  |
|  |  | mean | sum | std | mean | sum | std |  |  |  | lower | upper |
| control | 20 | 0.10 | 2 | 0.31 | 0.40 | 8 | 0.75 |  |  |  |  |  |
| intervention | 20 | 0.65 | 13 | 1.27 | 0.65 | 13 | 1.14 |  | 0.07 | 0.779 | -0.43 | 0.57 |
|  |  | E2. Practice Pages |  |  |  |  |  |  |  |  |  |  |
| control |  | 0.45 | 9 | 1.00 | 0.60 | 12 | 1.14 |  |  |  |  |  |
| intervention |  | 2.20 | 44 | 4.21 | 1.00 | 20 | 1.72 |  | -0.20 | 0.517 | -0.84 | 0.43 |
| Alert email sign-ups |  |  |  |  |  |  |  |  |  |  |  |  |
| group |  | list size (100k) |  |  | E3. CCG alerts |  |  |  |  |  |  |  |
|  |  |  |  |  | Before |  | After |  | Effect size | p value | 95% CI |  |
|  |  | mean | std |  | mean | std | mean | std |  |  | lower | upper |
| control |  | 2.57 | 1.20 |  | 1.20 | 1.67 | 0.15 | 0.37 |  |  |  |  |
| intervention |  | 3.34 | 1.56 |  | 0.85 | 1.31 | 0.35 | 0.59 | 0.36 | 0.077 | -0.03 | 0.61 |
|  |  |  |  |  | E4. Practice alerts |  |  |  |  |  |  |  |
| control |  | 2.57 | 1.20 |  | 0.95 | 1.64 | 0.25 | 0.55 |  |  |  |  |
| intervention |  | 3.34 | 1.56 |  | 2.90 | 3.75 | 0.45 | 1.00 | -0.07 | 0.790 | -0.59 | 0.45 |

Number of page views over one month on CCG pages showing low-priority measures (E1), and on equivalent practice pages (E2), for CCGs in intervention and control groups. Number of sign ups for CCG (E3) and practice (E4) OpenPrescribing email alerts, for CCGs in intervention and control groups.

CCGs in the intervention group on average set up 0.35 new email alerts in the three months following their intervention, compared to 0.15 in the control group. Taking into account baseline levels and CCG size to correct for the potential scope for new sign-ups, this did not quite meet the p= 0.05 threshold (p=0.077, coefficient 0.359; 95% CI −0.033 to 0.605). More practice alerts for constituent practices were also added in the intervention versus the control group (0.45 vs 0.25) but this difference was not close to being significant (p=0.790).

## Discussion

### Summary

An intervention delivered to CCGs by NHS England was successfully adapted into an RCT format, in a short timescale (under six months) and at low cost. We found no evidence that the intervention influenced low-priority prescribing. There was some possible evidence of increased engagement with data in the form of CCG-level email alert sign-ups.

### Strengths and weaknesses

This was an agile evaluation using an opportunistic RCT to provide a robust assessment of impact for an intervention that was already planned by NHS England. Key weaknesses relate to the low response rate and low numbers in our engagement outcomes. Eight of 20 invited CCGs did not respond to the invitation, despite being contacted again, and for one additional CCG no visit could be scheduled within the time frame. It is possible that these relatively small cost-saving opportunities were not seen as a priority over other ongoing work. There were some complexities around joint working between CCG MO teams which necessitated some grouping of CCGs in allocation and in delivery of the intervention. Not all joint team information was available prior to invitations being sent, but we gathered further information from responses to invitations and ascertained that two intervention CCGs would be visited together, and six additional CCGs from the non-allocated group were also included in the intervention. Our page views data in the engagement outcomes contained some very low numbers which made any meaningful analysis difficult. OpenPrescribing is a widely used site, and it was not practical to supply unique trackable links in this trial, so page views data may have been influenced by other factors and any changes would not necessarily be solely attributable to the impact of the intervention. There may have been other prescribing data tools used by CCGs and practices in response to the intervention which we could not measure. Due to resource issues the manual data collection necessary to assess our planned outcome on CCG workplans was not carried out; this might have indicated whether some CCGs did take on the action recommended in the intervention. However, the ultimate impact of this should have been a change in prescribing which we did not detect.

### Findings in Context

This one-off intervention delivered to CCGs medicines optimisation teams had no measurable impact on prescribing. The effectiveness of the intervention could have been limited by the specific issue addressed, NHS England low-priority products, the list of which had been released for over a year and already included as a policy for many CCGs for the period covered. In a retrospective study we previously found evidence that CCGs taking action on national antibiotic prescribing guidance did influence GP prescribing behaviour compared to those not taking action [9]. However, with limited capacity, CCG medicines optimisation teams have to balance competing priorities [10], and other issues may have provided greater opportunities for cost savings or improving patient safety. Further, reviewing patients on these low-priority products may not be prioritised by busy GPs, regardless of incentives from their CCG, particularly as many of the products are considered safe. However, a natural reduction likely occurs as fewer new scripts are initiated. We saw a general reduction in our prescribing measures across both groups, indicating that such a reduction was occurring, in line with previous trends [2]. Without stronger national action such as blacklisting or prior approval to prevent unnecessary prescribing [11], the gradual reduction is likely to continue until a plateau is reached. It is unclear whether these findings would be generalisable to other issues within prescribing, or to other CCGs outside of those which received the intervention.

It is difficult to identify any similar CCG (or ICB / sub-ICB Location) based interventions tested in an RCT, or at all. The impact of national interventions, such as guidelines and financial incentives, are typically only assessed retrospectively, making the effects difficult to confidently discern from other influences. When national programmes aimed at making cost savings in the NHS are assessed by their national bodies the methodology is not always accessible, making the findings difficult to scrutinise. Typically, interventions to influence prescribing behaviour that are tested by RCTs target practices directly rather than regional prescribing organisations/advisers [12], such as our previous RCT, which had a small impact on engagement with data but no significant impact on prescribing [13].

### Future Research

This trial demonstrates how routine interventions planned to improve uptake or adherence to healthcare guidance can be delivered as low-cost randomised trials to robustly assess their effectiveness. We estimate the total cost burden of this trial, in addition to the cost of the planned intervention, to cover seven days of senior input and 30 days more junior (Table S3). We propose that research funders or healthcare delivery organisations bodies consider developing a small team who can develop reproducible methods and frameworks to support agile evaluations of many healthcare interventions. Having established this workflow, the associated costs could be reduced in the future, with huge potential gains in the quality of evaluations available to healthcare decision makers.

## Conclusions

By assessing a healthcare intervention using a low-cost RCT, we found no evidence that a simple educational intervention delivered to CCGs had a measurable impact on prescribing behaviour. However our methods demonstrate that it is possible to deliver low cost RCT evaluations of the effectiveness of complex policy issues.

## Supporting information

Supplemental All

## Abbreviations

CCG: Clinical Commissioning Group
MO: Medicines Optimisation
NHS: National Health Service

## Declarations

### Ethics approval and consent to participate

This study was classified as service evaluation by the Oxford University Medical Sciences Interdivisional Research Ethics Committee; the need for ethical approval and informed consent was waived. Participants consented to receive the intervention, but were not aware they were in a trial. This study was non-medical, conducted among professional staff and using anonymised data, so the Declaration of Helsinki does not apply, but we followed best research practice.

### Consent for publication

Not applicable

### Availability of data and materials

The dataset(s) supporting the conclusions of this article is(are) available in the low-priority-CCG-visit-RCT repository, https://github.com/ebmdatalab/low-priority-CCG-visit-RCT. This study uses open, publicly available data alongside website/email usage data. Processed data is available within the study repository along with all analysis code.

### Authors’ Contributions

BM BG HJC conceived the study with input from the wider team. BM and HJC designed the methods with input from AW SB RP BR. HJC collected and analysed the data. All authors confirm that they had full access to all the data in the study and accept responsibility to submit for publication. HJC and BM drafted the manuscript. All authors contributed to and approved the final manuscript. HJC and BM contributed equally.

## Acknowledgements

We thank all participants involved in the intervention. Additionally we are grateful for the support of NHS Clinical Commissioners, NHS England’s Medicines and Policy team and the office of the Chief Pharmaceutical Officer.

