## Supplemental All for "Educational interventions delivered to prescribing advisers to influence primary care prescribing: a very low-cost pragmatic randomised trial using routine data from OpenPrescribing.net"

### SUPPLEMENTARY INFORMATION

***Note 1*** **- Changes to data extraction and analysis scripts**

- An error was found in import of “herbal” measure into the original analysis - was not correctly filtered. This was corrected for the final analysis. This would not have affected our ascertainment of eligibility as this measure was not available at the time of randomisation.
- We changed the data source used for practice-CCG allocations, due to broad changes in membership since the end of the trial with many CCGs merging. Instead of using the live/updated ‘practices` table, we used the *epraccur* table published October 2019 (NHS organisation data service (ODS)) to ensure that practices were aligned to the CCGs they were members of during the follow-up period.
- For similar reasons we made minor amendments around the source of alerts sign-up data - this was retrieved from archived copies from the end of the follow-up period (September 2019) rather than the latest data, to reduce the impact of changes to CCGs that would have caused alerts to be removed as users converted their alerts to the new organisations.
- We reverted to using a manually downloaded file instead of live linkage to Google Analytics service for page views data.

***Table S1.*** *NHS England commissioning guidance on 18 low priority treatments (NHS England 2017). †A measure on herbal remedies was not available through OpenPrescribing at the time of randomisation due to complexity of identification of relevant products, but was included in the intervention and in relevant outcomes.*

| Item | Rationale for inclusion in guidance |
|---|---|
| Co-proxamol | Items of low clinical effectiveness, where there is a lack of robust evidence of clinical effectiveness or there are significant safety concerns. |
| Dosulepin | Items of low clinical effectiveness, where there is a lack of robust evidence of clinical effectiveness or there are significant safety concerns. |
| Doxazosin prolonged release | Items which are clinically effective but where more cost-effective items are available, including items that have been subject to excessive price inflation. |

| | |
|---|---|
| Fentanyl Immediate-Release | Items which are clinically effective but where more cost-effective items are available, including items that have been subject to excessive price inflation. |
| Glucosamine and Chondroitin | Items of low clinical effectiveness, where there is a lack of robust evidence of clinical effectiveness or there are significant safety concerns. |
| Herbal treatments† | Items of low clinical effectiveness, where there is a lack of robust evidence of clinical effectiveness or there are significant safety concerns. |
| Homeopathy | Items of low clinical effectiveness, where there is a lack of robust evidence of clinical effectiveness or there are significant safety concerns. |
| Lidocaine Plasters | Items of low clinical effectiveness, where there is a lack of robust evidence of clinical effectiveness or there are significant safety concerns. |
| Liothyronine (including Armour Thyroid and liothyronine combination items) | Items which are clinically effective but where more cost-effective items are available, including items that have been subject to excessive price inflation. |
| Lutein and Antioxidants | Items of low clinical effectiveness, where there is a lack of robust evidence of clinical effectiveness or there are significant safety concerns. |
| Omega-3 Fatty Acid Compounds | Items of low clinical effectiveness, where there is a lack of robust evidence of clinical effectiveness or there are significant safety concerns. |
| Oxycodone and Naloxone Combination Product | Items of low clinical effectiveness, where there is a lack of robust evidence of clinical effectiveness or there are significant safety concerns. |
| Paracetamol and Tramadol Combination Product | Items which are clinically effective but where more cost-effective items are available, including items that have been subject to excessive price inflation. |
| Perindopril Arginine | Items which are clinically effective but where more cost-effective items are available, including items that have been subject to excessive price inflation. |
| Rubefacients (excluding topical NSAIDs) | Items of low clinical effectiveness, where there is |

| | |
|---|---|
| | a lack of robust evidence of clinical effectiveness or there are significant safety concerns. |
| Once Daily Tadalafil | Items which are clinically effective but where more cost-effective items are available, including items that have been subject to excessive price inflation. |
| Vaccines administered exclusively for the purposes of travel | Items which are clinically effective but due to the nature of the product, are deemed a low priority for NHS funding |
| Trimipramine | Items which are clinically effective but where more cost-effective items are available, including items that have been subject to excessive price inflation. |

***Table S2. Prescribing outcomes for sensitivity analysis.*** *CCGs which did not receive the intervention (n=9) were re-grouped into the control group. Outcomes replicate P1 and P2, i.e. total cost and number of items all 18 low-priority prescribed per 1000 population in the intervention versus control groups.*

| | | Cost per 1000 patients | | | | | | | | |
|---|---|---|---|---|---|---|---|---|---|---|
| | | Baseline | | Follow-up | | | | | 95% CI | |
| **allocation** | **count** | **mean** | **std** | **mean** | **std** | **Change** | **Effect size** | **p value** | **lower** | **upper** |
| **control** | 29 | 1,248.4 | 209.7 | 1,042.4 | 187.3 | -206.0 | | | | |
| **intervention** | 11 | 1,158.1 | 154.1 | 940.9 | 163.7 | -217.2 | -35.4 | 0.387 | -117.2 | 46.5 |
| | | **Items per 1000 patients** | | | | | | | | |
| **control** | 29 | 35.58 | 10.90 | 28.78 | 9.30 | -6.79 | | | | |
| **intervention** | 11 | 27.55 | 11.02 | 22.22 | 8.73 | -5.33 | -0.52 | 0.732 | -3.60 | 2.55 |

***Table S3*** *We estimated the additional costs involved in running the evaluation as an RCT. Costs were limited to person time.*

| **Role** | **Activities** | **Time (rounded to nearest day)** |
|---|---|---|
| Researcher (HC, AW) | Planning, ethics enquiries, protocol development, data extraction, analysis manuscript preparation | 30 |
| Researcher (HC) | Submission of protocol, registry entry and results, and submission of Academic manuscript activities (at time of submission) | 2 |
| Principal Investigator Support (BG) | Planning, supervision, analysis, detailed manuscript review | 2 |
| Senior statistician support (RP) | Planning, supervision, results review | 1 |
| Software engineer support (SB) | Code review and refactoring, analysis, results review | 1 |
| Intervention team (BMK, BR) | Planning, analysis, manuscript preparation | 2 (this does not include the cost of delivering the intervention, which was happening anyway, and outside the scope of this study) |
